# TRANS-AORTIC VALVULAR FLOW DYNAMICS AND LEFT VENTRICULAR EJECTION FRACTION FOR MONITORING RECOVERY OF PATIENTS WITH LEFT VENTRICULAR SYSTOLIC HEART FAILURE

**DOI:** 10.1101/2023.04.25.23289129

**Authors:** Viswajith S. Vasudevan, Keshava Rajagopal, Jesus E. Rame, James F. Antaki

## Abstract

Durable mechanical circulatory support in the form of left ventricular (LV) assist device (LVAD) therapy is increasingly considered in the context of the recovery of native cardiac function. Progressive improvement in LV function may facilitate LVAD explantation and a resultant reduction in device-related risk. However, ascertaining LV recovery remains a challenge. In this study, we investigated the use of trans-aortic valvular flow rate and trans-LVAD flow rate to assess native LV systolic function using a well-established lumped parameter model of the mechanically assisted LV with pre-existing systolic dysfunction. Trans Aortic Valvular Ejection Fraction (TAVEF) was specifically found to characterize the preload-independent contractility of the LV. It demonstrated excellent sensitivity to simulated pharmacodynamic stress tests and volume infusion tests. TAVEF may prove to be useful in the ascertainment of LV recovery in LVAD-supported LVs with pre-existing LV systolic dysfunction.

## Introduction

Left ventricular assist devices (LVADs) have been used to treat tens of thousands of patients with advanced heart failure (HF), most commonly as an alternative to heart transplantation (HT). Modern LVADs have improved survival, exceeding 85% after 1 year and 45% after 5 years^1,2^. However, even though short-term outcomes of LVAD implantation approximate those of HT, long-term outcomes are substantially inferior, largely due to time-accruing LVAD-related complications (including but not limited to: native aortic valve regurgitation, outflow graft complications, device thrombosis, pump mechanism malfunction, inflow cannula obstruction, bleeding complications, and infection). Accordingly, when native LV systolic function sufficiently improves, it is often desirable to explant the LVAD. Consequently, the prospects for using LVADs as a “bridge to recovery” has received increasing interest^3–9^. However, the reported incidence of post-LVAD implantation LV recovery remains low (4% at five years for adults after receiving an LVAD^2^ and 8.1% after 12 months for pediatrics^10^). In cases in which LV recovery is suspected, it is ideal to be able assess native LV systolic function independent of LVAD action. A systematic, objective approach would be to characterize the pressure-volume diagrams of the LV over a range of loading conditions – i.e., comprehensive Frank-Starling and Anrep mechanism assessments. This is challenging due to: (1) the continued MCS and LV unloading of the LVAD even at low speeds, and (2) unreliable *in vivo* measurement of trans-LVAD pressure difference (between LV and ascending thoracic aorta) and volumetric flow rate.. Standard practice involves so-called “ramp” and “turn-down” studies, in which the speed of the VAD is increased or decreased. Additional volume infusion studies further characterize the response to increased preload. However, none of these interventions has been shown to reliably predict what will happen after the LVAD is explanted.

Therefore, this study aimed to provide an approach to assessing native LV systolic function in the LVAD-supported circulation. The model distinguishes systemic arterial blood flow due to blood within the LV being ejected across the aortic valve, from systemic arterial blood flow due to blood within the LV flowing through the LVAD system. Our previous work demonstrated how this flow division is affected by pump speed^11^. We also introduced a new metric of LV contribution, TAVEF, the Trans-Aortic Valvular Ejection Fraction, which provides a measure of native LV systolic function function independent of LVAD contribution.

In this study, we employ a well-established *in silico* model of the cardiovascular system to simulate clinical *speed titration* and *volume infusion* tests for a range of HF states corresponding to respective NYHA classifications^12–14^, and exercise conditions. We present the correlation between,, LVEF, and the total output over the full range of pump speed to characterize the dynamics of the aortic valve and its role in assessing recovery. Finally, we investigate the efficacy of TAVEF as a surrogate for monitoring recovery and predicting post-LVAD explantation LV dynamics.

## Materials and Methods

### System of Systems Modeling of the Mechanically Assisted Circulation

This simulation study employed an established lumped parameter model of the mechanically assisted cardiovascular system with dilated heart failure^15^ incorporating the HeartMate 3^®^ VAD reported previously.^16^ This model can be considered as a *system of systems* (SoS) that comprises the Cardiac System (Heart), Systemic and Pulmonary Circulation Systems, and the VAD system, composed of a pump, inlet cannula, and outflow graft.^17^ (See Fig. 1.) The primary input variable to the model is the rotational speed of the LVAD (revolutions per minute, RPM). The main parameters defining the model are those representing the cardiac dynamics of the dilated ventricle, systemic and pulmonary resistances and compliances, and unstressed volume. The parameters of the unimodal elastance function are (i) the slopes of the ascending and descending limbs of the end-systolic pressure-volume relationship, (ii) the pressure axis intercept, (iii) the unstressed volume intercept, and (iv) end-diastolic pressure-volume relation (EDPVR) parameters. (See Supplemental Data 1 for description of the model and parameters.) Disturbances to the model are dosage of drug delivery and volume of blood infused.

**Fig. 1:**
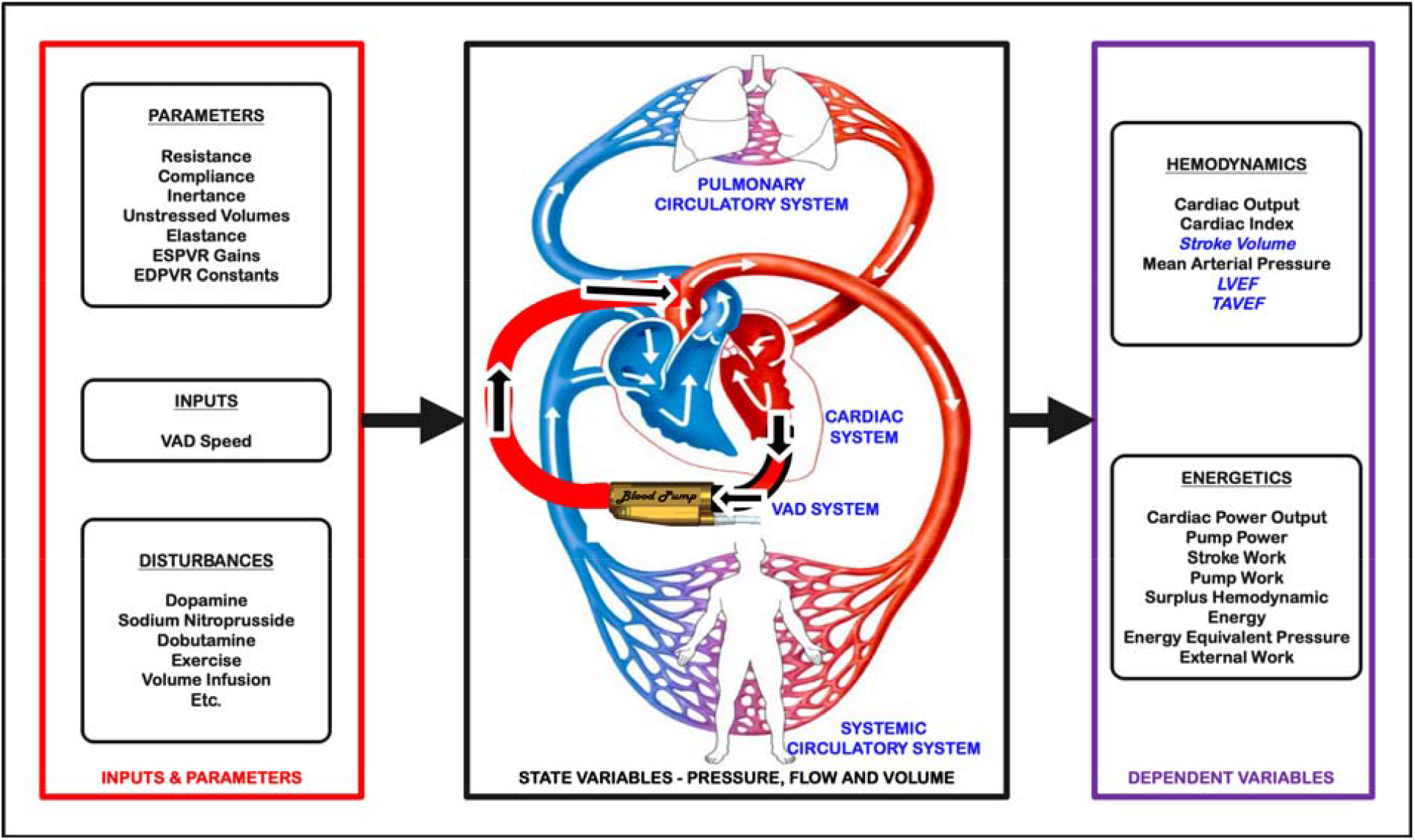
Representation of the Mechanical Support System of Systems (SoS) including the Cardiac, LVAD, Systemic Circulatory and Pulmonary Systems (inside the box). The variables describing the SoS, input & parameters, state and dependent variables are listed. The colored arrows inside the SoS box indicate the direction of flow of blood.

The state variables describing the SoS are the pressures, volumes, and flow rates of the constituent systems, specifically ventricular pressure and volume, arterial pressure, total cardiac output, pulmonary arterial and flow, and VAD head pressure and flow rate.

### Simulation of Heart Failure in terms of NYHA Classes

Different degrees of heart failure is commonly described clinically in terms of cardiac symptoms, categorized by New York Heart Association (NYHA) Classes from NYHA I to NYHA IV^12–14^. In this simulation, the model parameters were adapted to reproduce baseline hemodynamics for each of these classes of heart failure, reported previously by Ky et al.^18^, following the procedure outlined by Burkhoff^19^. (See *Table ST 1*.*1* in *Supplemental Data 1*.) In addition, two degrees of recovery were considered in this study derived from the definition employed in the RESTAGE clinical trial^9^ namely: *partial recovery* (LVEF between 45 and 59%) and *total recovery* (LVEF between 60 and 70%). The baseline hemodynamics for the six levels of cardiac function and the corresponding pressure-volume (PV) loops of the left ventricle are provided in *Supplemental Data 1, Table ST 1*.*2*, and *Figure SF 1*.*1*, respectively. These values are in good agreement with published data^18,19^.

### Simulated VAD Speed Turn-Down Study

A *turn-down study*, also known as *reverse ramp test*^20^, is performed by gradually reducing the rotational speed of the VAD in a controlled, supervised setting, and observing the hemodynamic response. Echocardiography provides observable evidence of ventricular dynamics, such as stroke volume, ejection fraction, aortic valve opening, and end-diastolic area/volume. The simulation was performed at successively decreasing speed from 9000 RPM to 0 RPM in increments of 1000 RPM. Thereafter, removal of the VAD was simulated to provide “ground truth” assessment of cardiac function. For each condition, the simulation was permitted to reach a steady state before calculating the dependent variables.

### Pharmacodynamic Challenge

Sodium nitroprusside and dopamine are two drugs used to augment left ventricular output (the former via afterload reduction, the latter via contractility increase), providing a better assessment of the pumping capacity of the heart. In the simulation, we perturb affected parameters of the model as described by Gopinath et al.^21^ summarized in Table 1.

**Table 1:**
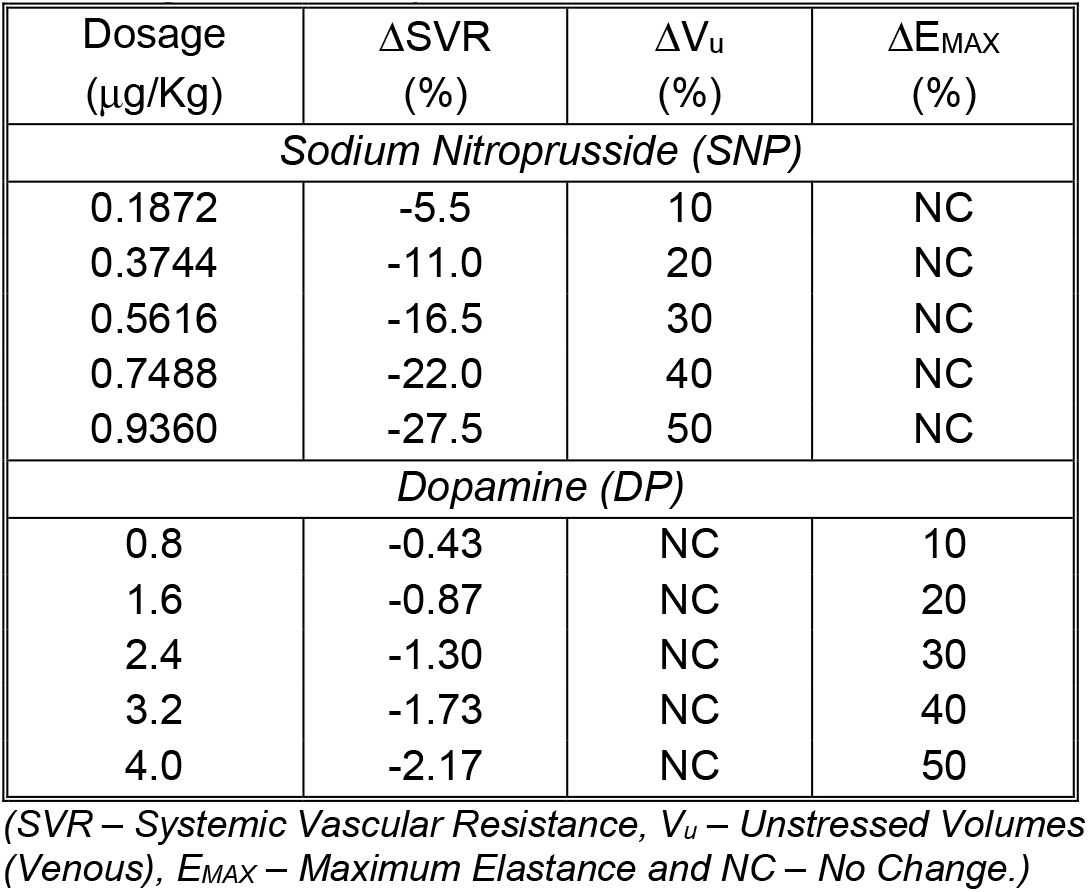
Dosage of Pharmacologic Drugs and Equivalent Changes (%) in Lumped Parameters Used in Simulation.

| Dosage<br>($\mu\text{g/Kg}$) | $\Delta\text{SVR}$<br>(%) | $\Delta V_u$<br>(%) | $\Delta E_{\text{MAX}}$<br>(%) |
| --- | --- | --- | --- |
| *Sodium Nitroprusside (SNP)* |  |  |  |
| 0.1872 | -5.5 | 10 | NC |
| 0.3744 | -11.0 | 20 | NC |
| 0.5616 | -16.5 | 30 | NC |
| 0.7488 | -22.0 | 40 | NC |
| 0.9360 | -27.5 | 50 | NC |
| *Dopamine (DP)* |  |  |  |
| 0.8 | -0.43 | NC | 10 |
| 1.6 | -0.87 | NC | 20 |
| 2.4 | -1.30 | NC | 30 |
| 3.2 | -1.73 | NC | 40 |
| 4.0 | -2.17 | NC | 50 |

(SVR – Systemic Vascular Resistance, $V_u$ – Unstressed Volumes (Venous), $E_{\text{MAX}}$ – Maximum Elastance and NC – No Change.)

### Volume Infusion Study

Another clinical intervention that may be performed to assess preload reserve of the heart is to infuse fluid^22,23^. This was simulated by adjusting the initial conditions of total blood volume from a baseline value of 5300 ml^24,25^ to 5800ml in in steps of 100 ml.

## RECOVERY ASSESSMENT USING SOS – DEPENDENT VARIABLES

The dependent variables employed to assess ventricular function, hence recovery, are derived from the state variables of the model and include stroke volume (SV), LVEF, and TAVEF as defined in our previous work^11,26^. The left ventricular stroke volume, SVLV, is defined using the traditional formula as the difference between end-diastolic and end-systolic ventricular volume, EDV, ESV, respectively:

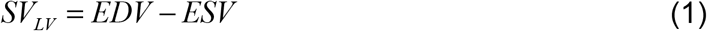

In the assisted circulation, a portion of this blood volume is ejected through the aortic valve, and the remainder is routed through the VAD into the systemic circulation. Accordingly, the *trans-aortic-valve stroke volume*, SV_AV_ is defined as the net volume of blood introduced into the systemic circulation through the aortic valve during one cardiac cycle,

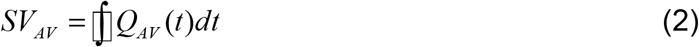

the *trans-VAD stroke volume*, SV_AV_ is defined as the net volume of blood introduced into the systemic circulation through the VAD during one cardiac cycle.

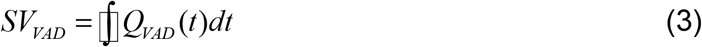

and the *total stroke volume*, SVT is the sum:

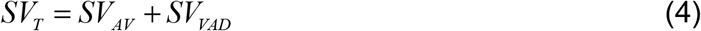

In addition, the Trans-Aortic Valvular Ejection Fraction, TAVEF is defined as the relative ejection fraction corresponding to SV_AV_:

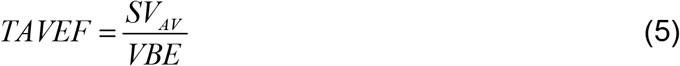

where VBE is left ventricular volume at the start of the ejection phase of the cardiac cycle. The value of TAVEF is equivalent to LVEF in the absence of a VAD. It reduces with increasing VAD support, reaching a minimum of zero when the aortic valve ceases to open.

## Results

The present study considered relationship of the heart and the VAD as a system of systems (SoS) with VAD speed as an input, and revealed a fascinating interaction. At greater speed, and the aortic valve is closed, there is cooperation between LV and VAD.

### TURN DOWN STUDY: DISTRIBUTION OF STROKE VOLUME BETWEEN VAD AND AORTIC VALVE

Fig. 2 plots SV_AV_, SV_VAD_, and SV_T_ for simulated turn-down studies under heart failure conditions corresponding to NYHA I and IV. Significant features observed from the plot are:

**Fig. 2:**
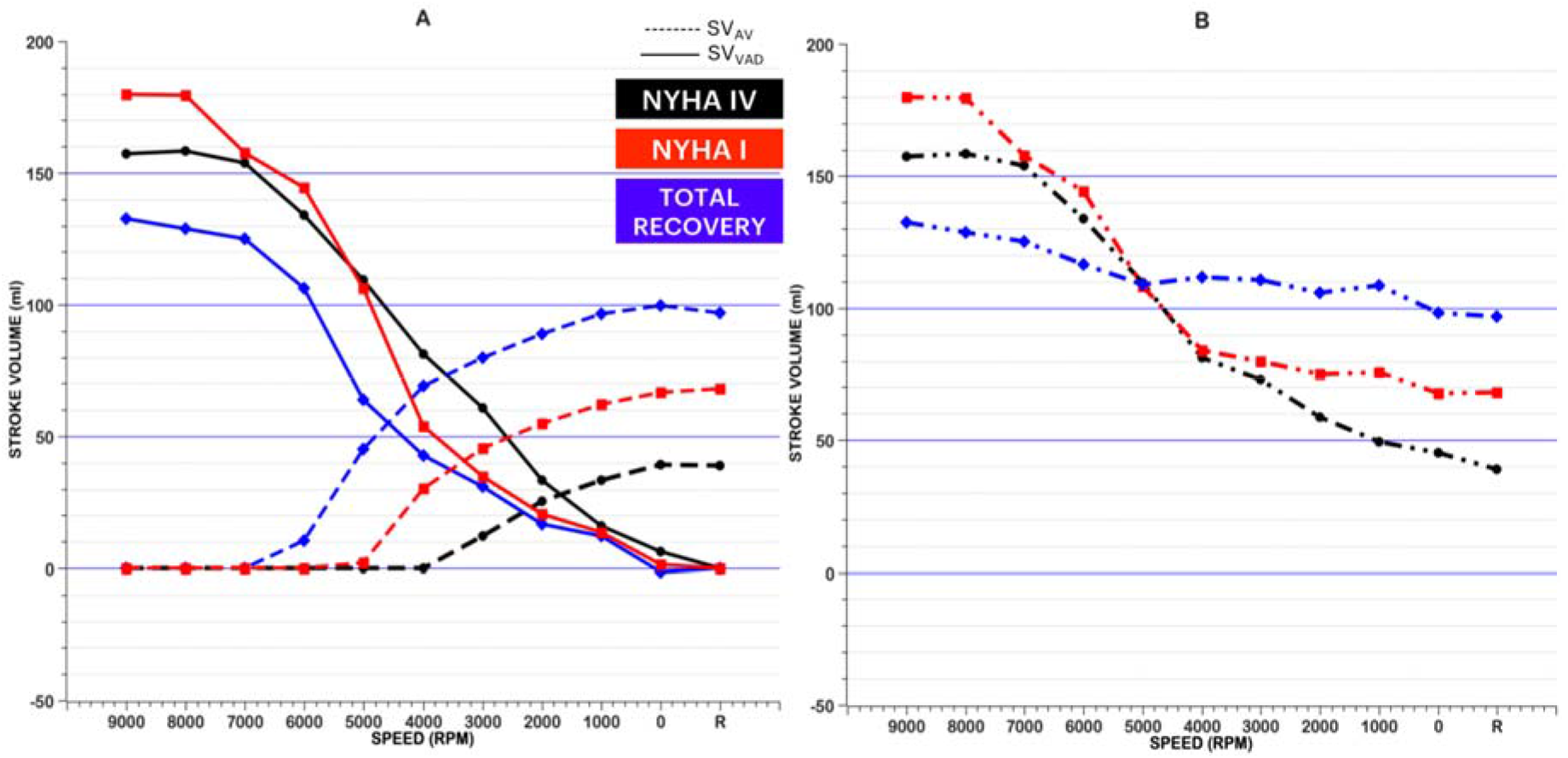
Simulated turn down study. (A) SV AV, stroke volume through the aortic valve, and SVVAD, stroke volume through the VAD and (B) SVT, the total stroke volume for NYHA IV, NYHA I and Total Recovery cases as a function of VAD Speed.

I.Initially, SVVAD is equivalent to SVT at greater speeds for all classes, as the aortic valve remains closed (SV_AV_ = 0).

II.Reducing speed below 6000 RPM for a recovered ventricle (5000 for NYHA I and 4000 for NYHA IV), the aortic valve opens, and SV_T_ exceeds SV_VAD_.

III.With further speed reduction, the proportion of stroke volume through the aortic valve (SV_AV_) exceeds the stroke volume through the VAD (SV_VAD_). This reversal occurs at 4600 RPM for Total Recovery, 3200 RPM for NYHA I and 1700 for NYHA IV.

IV. As speed approaches zero, SVVAD becomes zero or slightly negative.

V. For speed above 5200 RPM, SV_VAD_ for NYHA I exceeds NYHA IV; for speeds below 5200 RPM, SV_VAD_ for NYHA I is less than NYHA IV. (Note: For total recovery, SV_VAD_ is lower for all speeds.)

VI. For all speeds, the SVT for NYHA I is greater than or equal to SV_T_ for NYHA IV.

VII.At lower speeds SVT is directly related to cardiac contractility: greatest for Total Recovery, lowest for NYHA IV.

The specific nature of different speed titration responsiveness of SV_VAD_ in profiles of Total Recovery, NYHA IV and NYHA I HF, in Fig. 2, is perhaps not what would be predicted, in that clean trends depending upon HF disease severity were not found. The reasons for this are not entirely clear, but seem to be related at least in part to the incorporation of baroreflex into the model. The following discussion tries to explain from the studies results the trend observed.

The baroreflex response to the different heart condition and hence its effects on the Heart Rate, play a major role in affecting the SV_VAD_. Fig. 3 shows the plot of the T as a function of heart condition and speed. During ventricular suction, the Heart Rate is suppressed the highest for NYHA IV followed by NYHA I and Total Recovery. Thus, the cardiac cycle, T, is varied inversely. Meaning the cardiac cycle for NYHA IV is greatest followed by NYHA I and Total Recovery. As defined above, the volume of blood flowing through the VAD over a cardiac cycle is the integral of the product of flow and change in time. Thus, for the stroke volume, the value is dependent on the area under the VAD flow curve in each cycle considered. The effect of heart condition on the metrics of the flow rate signal (peak-to-peak and maximum value) is less pronounced than the change in cardiac cycle. Physiologically we might not be getting reliable volume calculations at speeds >6000 RPM due to suction, thus going by the math behind the simulations, we see the trend where SVVAD for NYHA I > NYHA IV > Total Recovery for speeds >6000 RPM. Between 6000RPM and 7000RPM, the volume for both NYHA I and IV are almost similar in this case. For speeds below 6000 RPM, the influence of the VAD is greater and thus the flow rate signal has a peak-to-peak for NYHA IV > NYHA I > Total Recovery, with the minimum value having greater negative flow at Total Recovery > NYHA I > NYHA IV. Thus, given heart rate and the flow signal, the stroke volume through the VAD follows, NYHA IV > NYHA I > Total Recovery.

**Fig. 3:**
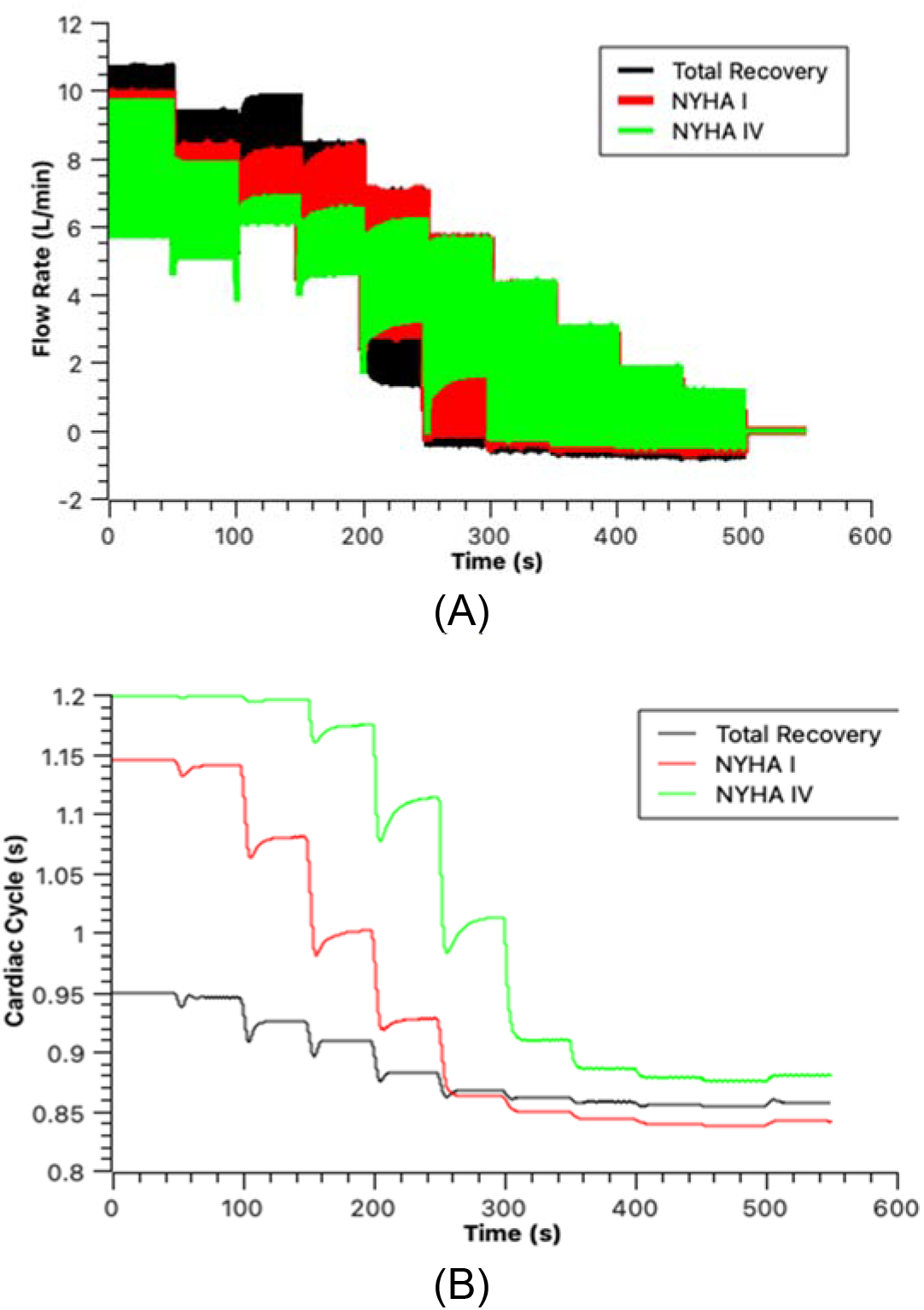
(A) Q _VAD_ vs Time for turn down study in the three cases (NYHA IV, NYHA I and Total Recovery). (B) Duration of Cardiac Cycle vs Time for change in speed for turn down study in the three cases (NYHA IV, NYHA I and Total Recovery).

Another observation the study made is the absence of or minimal retrograde flow, in the VAD, reflected by a minimal negative volume. The inertance of the outflow graft is observed to play a role in adjusting the retrograde flow occurring. The study tested the hypothesis that greater the inertance value, the lesser the retrograde flow. Fig. 4, highlights two example cases that includes a high value (order of 1/10) for the inertance value and a ∼0 inertance value. It is clearly observed that higher the inertance value, the retrograde flow is reduced but does not go to 0. Whereas while we calculate the stroke volume, depending on the negative flow canceling out the positive flow while performing the numerical integration, it might provide a close to 0 ml net volume, which is the observation the study makes in point IV above.

**Fig. 4:**
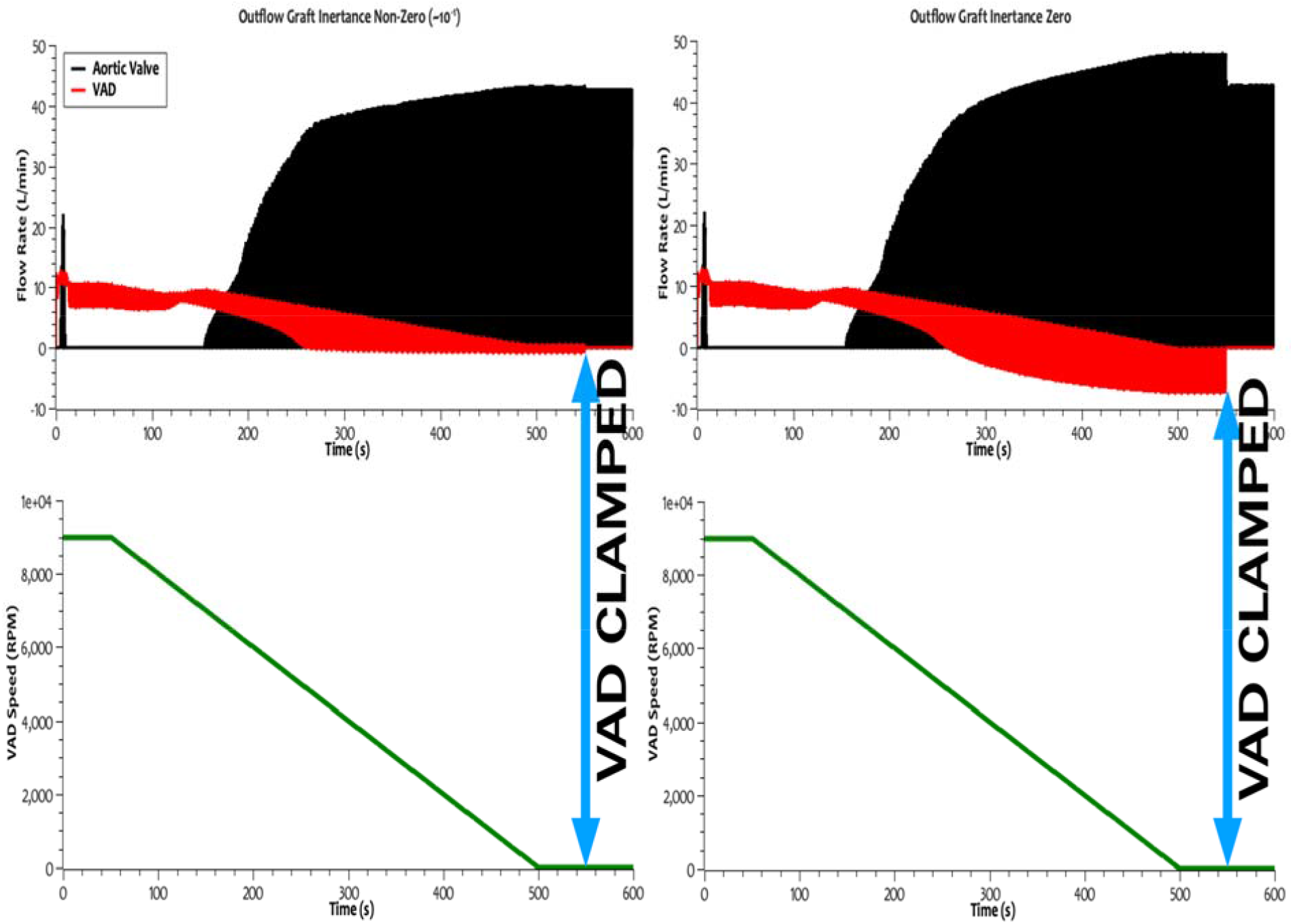
Effect of Outflow Graft Inertance on Trans-Aortic Valvular and Trans-VAD flows as seen during the turn-down study. The turn-down begins at t = 50sec and at t = 550secs, the clamping of the VAD is simulated.

### Response LVEF and TAVEF to Reduced VAD Speed

Ejection fraction measures, LVEF and TAVEF, provide a better understanding of the complimentary, and counteracting effect of the heart and the VAD. Fig. 5(A) plots the values of TAVEF as a function of speed for the simulated turn-down study. The aortic valve is closed at speeds above 7000 RPM; hence, TAVEF is 0. TAVEF becomes nonzero below 5000 RPM for NYHA I and below 4000 RPM for NYHA II; all classes have a nonzero TAVEF at speeds below 3000 RPM. For reference, the figures indicate the 45% EF criterion specified by the RE-STAGE study for recovery^9^.

**Fig. 5:**
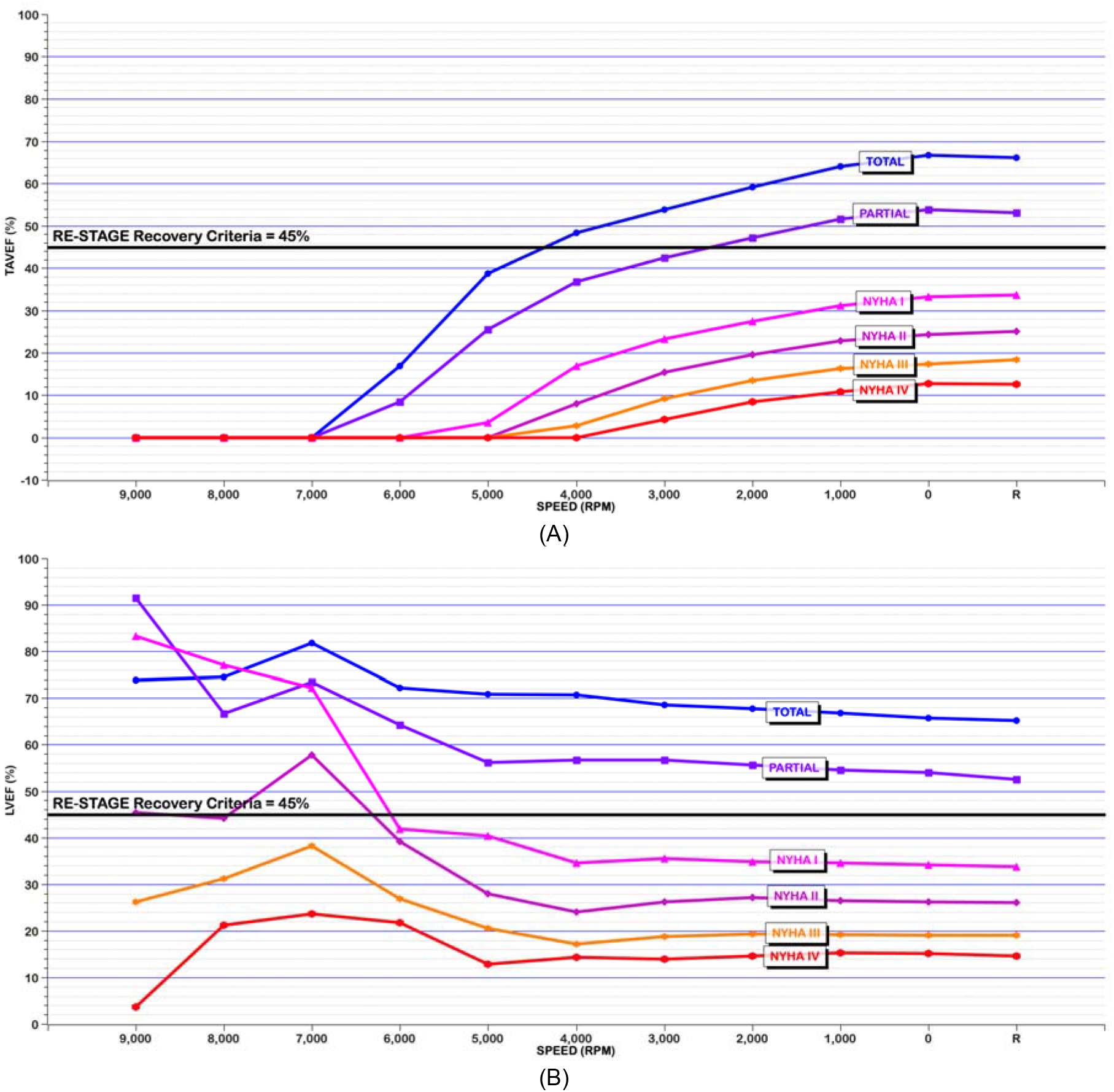
(A) TAVEF for the different cases in a turn-down study (B) LVEF for the different cases in a turn-down study.

Compared to TAVEF, total LVEF is less sensitive to turn-down intervention. For example, LVEF is nonzero for all speeds and conditions of the heart. As an illustration, if one considers the specific case NYHA III (orange lines in the graph) for speed from 5000 to 2000 RPM, LVEF reduces from 22% to 20.5%, whereas TAVEF increases from 0 to 11%. This insensitivity of LVEF to speed is due to the counteracting contribution of the heart (SV_AV_), which increases with decreasing speed, while SV_VAD_, decreases. In general, for all NYHA classes, the compensatory effect between the heart and VAD maintains LVEF almost constant for all levels of VAD support. From the perspective of myocardial recovery, greater ventricular contractility “limits” the contribution of the VAD. Consequently, TAVEF is a more effective indicator of recovery. Another interesting observation is that TAVEF for the case simulated for removal of VAD is equal to those at a speed of 1000 RPM.

Performing turn-down studies especially in patients who demonstrated recovery or reverse modeling, it has been observed LVEF to always increase with reduction of VAD Speed. In comparison, the trend in Fig. 5(B), an important factor in overall LVEF decreasing slightly is likely due to the unimodal ESPVR used in the study alongwith the VAD speed reduction induced LV distension^11,15^.

Additionally, if the LV had previously been “over-underloaded”, i.e., run at speed too high to facilitate recovery, that a drop in LVEF would not be observed. Clinically, it is probably more likely to see this latter scenario in the recovering LV, because LV reverse remodeling would have occurred, and LVEDV would be lower.

Interestingly, the effect of the inertance of the outflow graft on LVEF or TAVEF seems to be minimal. The increased retrograde flow increases the end-diastolic and end-systolic volumes and affects the values of LVEF and TAVEF minimally. (higher retrograde flow LVEF at Clamped 38.3%, TAVEF at Clamped 38.3%, lower retrograde flows LVEF at Clamped 36.7% andf TAVEF is also 36.7%) These values are only for this specific example, and the trend is certainly observed for all the examples considered.

## Simulation of Pharmacodynamic Interventions

Fig. 6(A) and (B) show the effect of sodium nitroprusside (SNP) and dopamine (DP) challenge on TAVEF, respectively, as a function of VAD speed. For speeds above 7000 RPM, TAVEF is 0%, irrespective the adrenergic or vasoactive effects of SNP and DP.

**Fig. 6:**
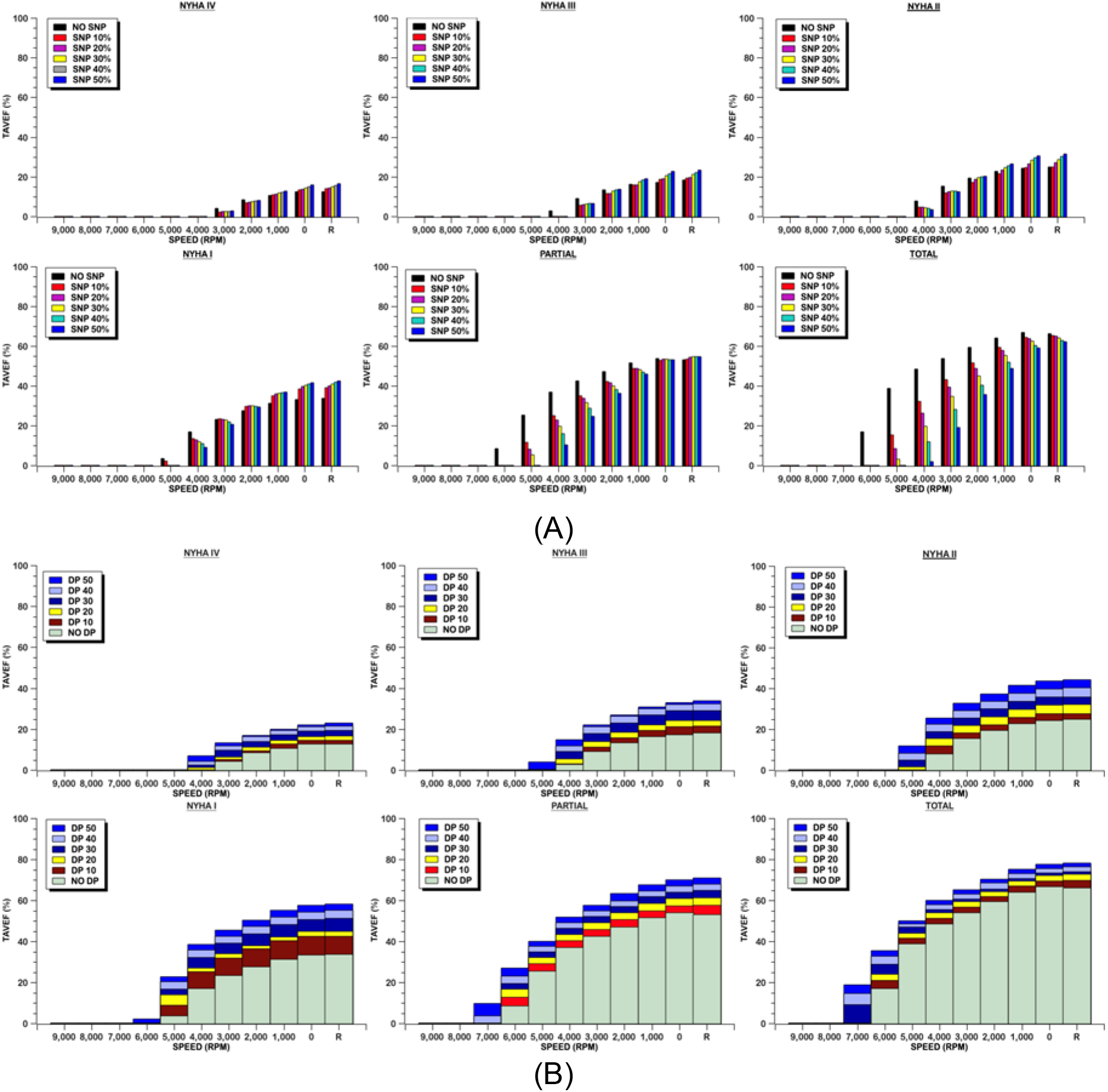
TAVEF values for (A) Sodium nitroprusside (SNP) infusion at 10% - 50% changes in the parameters affected by SNP. (B) Dopamine (DP) addition resulting in 10% - 50% change in parameters. (Horizontal line indicates 45% EF criterion. R refers to VAD Removal.)

### TAVEF and SNP

The effect of SNP on TAVEF is seen to be a function of speed, cardiac function, and RPM. For more severe classes of heart failure, increasing dosage of SNP has a positive effect on TAVEF at lower speeds, and simulated recovery. One exception is an initial drop in TAVEF with initial dose of SNP in the range of speed from approximately 3000 to 4000 RPM. As cardiac function improves (from NYHA III to II to recovery) the positive response of TAVEF to SNP becomes less pronounced and eventually becomes negative. This can be explained by decreased ventricular preload due to translocation of blood to the venous reservoir which has a more pronounced effect in the health heart. The effect of SNP on LVEF is summarized in Supplemental Data 2.

### TAVEF and DP

The change in TAVEF with DP is more consistent across the different types of heart conditions and speeds of VAD. (See Fig. 4B.) TAVEF increases with dosage of DP and improved heart conditions at all speeds in which TAVEF is nonzero. This response is due to increased ventricular contractility and a decreased vascular resistance. The effect of DP on LVEF is also summarized in Supplemental Data 2.

## Simulation of Volume Infusion

The effect of 100 ml increments of blood infusion on TAVEF and LVEF is illustrated in Fig. 7 for two representative speeds of 3000 RPM and 6000 RPM. The former is chosen as it provides partial support for all cardiac conditions studied; the latter is chosen because it provides full support for all NYHA classes and provides partial support in the two recovery conditions. At 3000 RPM the effect of infusion on both TAVEF and LVEF is negligible for all four failure conditions studied. There is a slight positive effect observed in TAVEF, but not LVEF, for the two recovery conditions. At 6000 RPM, TAVEF is 0% for all classes of heart failure, but demonstrates a positive response to infusion for both recovery conditions. In contradistinction, the effect of volume infusion on LVEF was minimal.

**Fig. 7:**
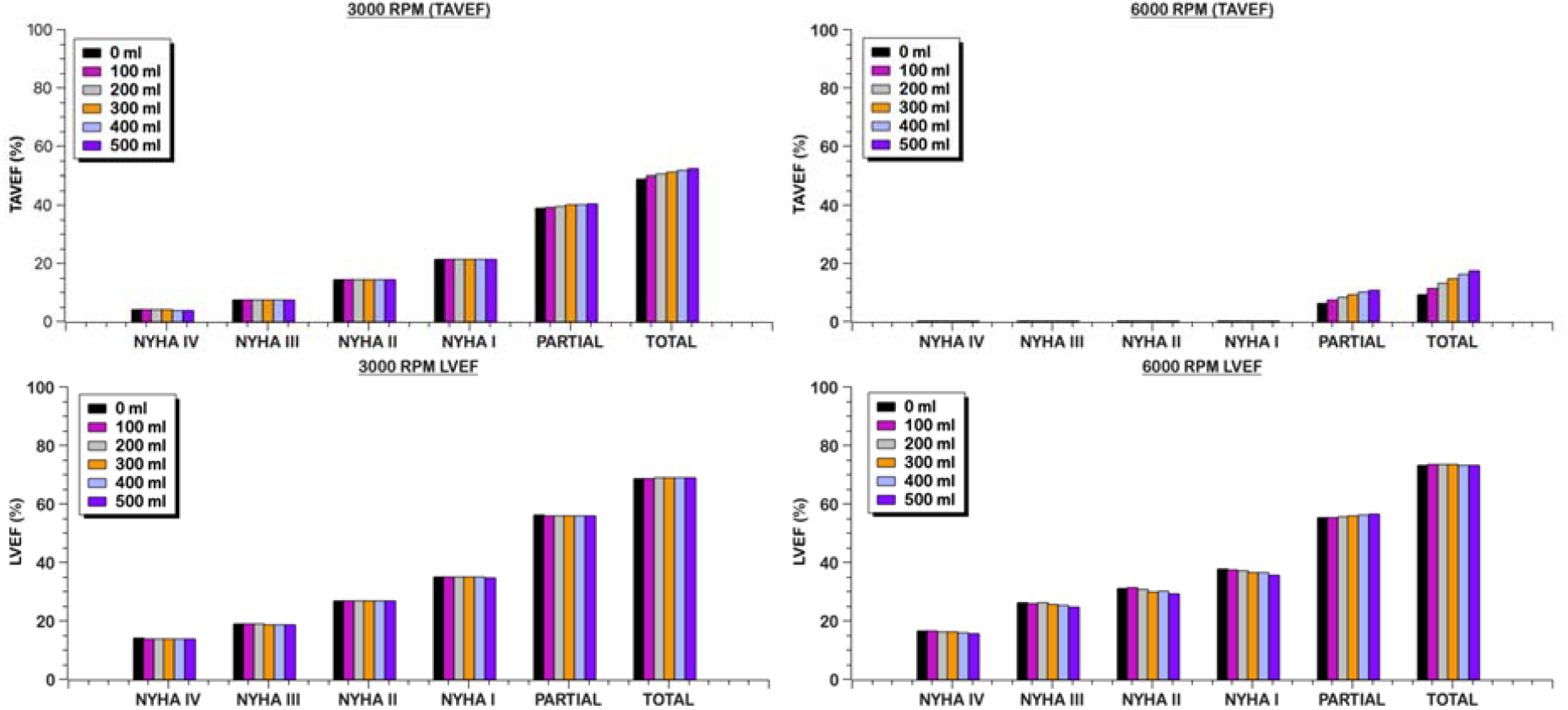
TAVEF and EF for increments of 100 ml of blood volume infusion at (A) 3000 RPM and (b) 6000 RPM

## DISCUSSION

The prospect of VAD therapy as a bridge-to-recovery, when possible, is a desirable outcome for many heart failure patients. In clinical practice, this requires a protocol to evaluate, or prognosticate, cardiac function independent of the VAD. A common practice is the so-called *turn-down* study, which attempts to reveal the pumping capacity of the native left ventricle by gradually reducing the support offered by the VAD. Interventions such as dopamine challenge and volume infusion provide additional insight to the condition of myocardium and preload-response of the ventricle.

Our simulations of the mechanically assisted circulation indicate that LVEF is insensitive to changes in VAD speed below 5000 RPM (for HeartMate III). However, LVEF measured at VAD speed of approximately 5000 is virtually equivalent to LVEF at removal of the VAD, for the range of cardiac function studied here. LVEF shows a one-to-one correspondence to the improvement of cardiac function. However, there are instances (increased regurgitation of blood in the VAD due to insufficient outflow graft inertance or ventricular suction) when LVEF could not be a representative measure for judging recovery, due to the way SVLV is defined. An instance of such insensitivity to change in physiology is seen in the current study with the volume infusion discussed above, where LVEF does not exhibit much of a change to increasing volume levels. The change reflected in SV_LV_ is negligible as compared to change in EDV.

Therefore, we introduced an alternate metric, TAVEF, which provides a more sensitive indicator of the native function with minimal perturbation of VAD speed. For example, Fig. 4 illustrates that, for the partial recovery, decrementing speed by 1000 RPM from a nominal operating speed of 6000 RPM effects an increase TAVEF from 8% to 25% (+17%), whereas LVEF decreases from 64% to 56% (−8%). Intuitively as the support provided by the VAD decreases, the share of perfusion from the ventricle is expected to increase, which in this current study as seen in Fig. 5, LVEF shows either a decrease or plateaus. TAVEF is a measure that is devoid of the effect of the VAD, and hence can reflect exactly the response to the support provided by the VAD. And since TAVEF considers the outflow through the aortic valve, the changes in the instantaneous geometry of the ventricle can be overcome, thereby eliminating issues such as regurgitation of blood through the VAD and ventricular suction (TAVEF = 0).

An illustrative example is provided in Fig. 8 that considers successive turn down studies of a hypothetical HeartMate III patient over the course of 360 days. At POD 1 a decrement of speed from 6000 to 3000 RPM results in a decrease in LVEF from 22 to 14% and increase in TAVEF from 0% to 4%. At a later date (POD 60), simulating moderate improvement from NYHA IV to NYHA III, LVEF responds to same speed turn-down by decreasing from 27 to 19% while TAVEF increases from 0% to 9%. At POD 360, simulating partial recovery, LVEF reduces from 64 to 57%, and TAVEF increases from 8% to 42%. Although both LVEF and TAVEF provide a positive correlation with contractility (recovery), TAVEF is much more responsive to pharmacological intervention. At all POD days considered, TAVEF decreases with SNP introduction and increases with DP, whereas LVEF shows little to no change with SNP and a slight increase with DP.

**Fig. 8:**
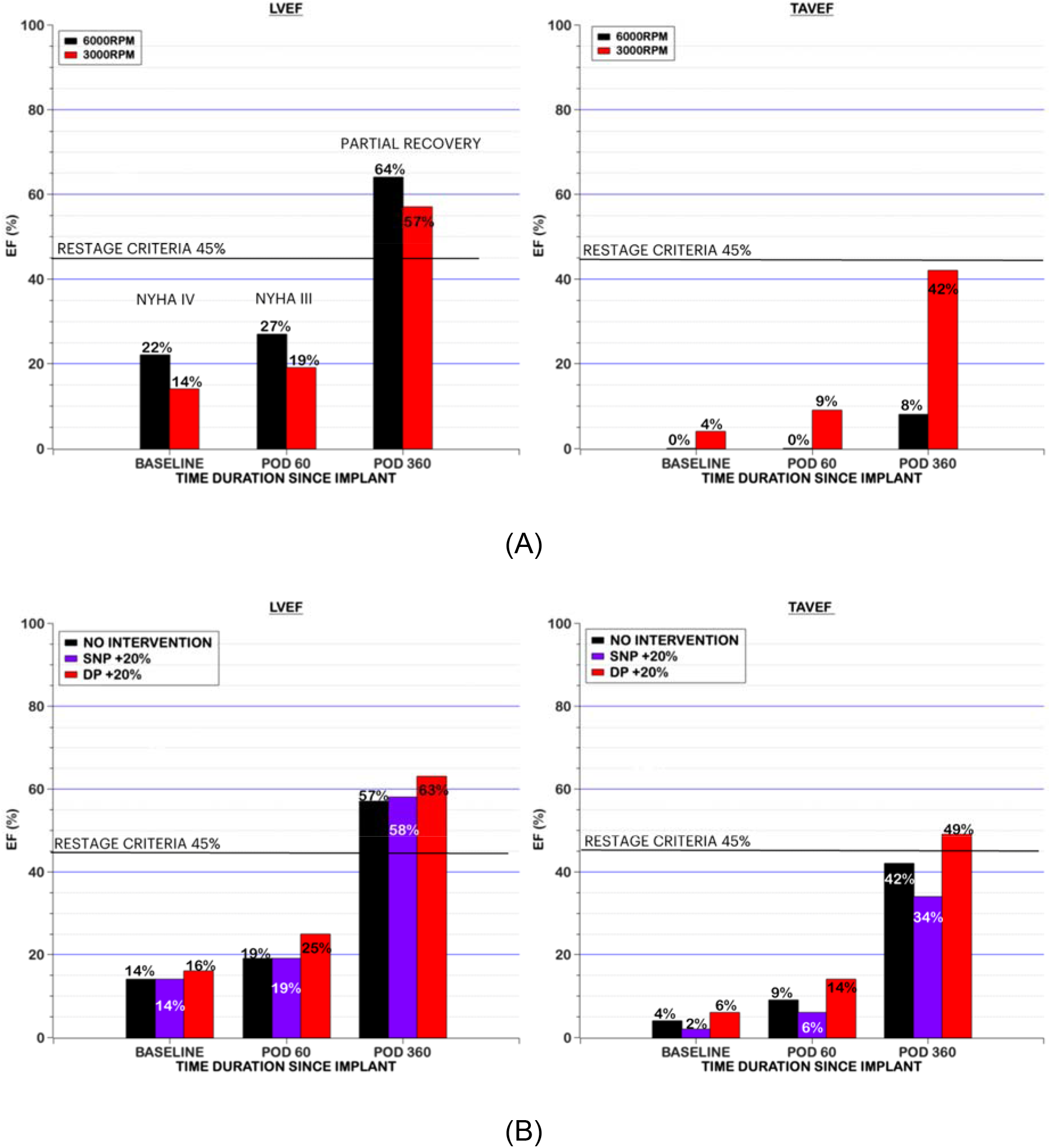
Illustrative example for assessment of recovery in a hypothetical patient over the course of 360 days. Timeline variations in TAVEF and LVEF are provided for (A) speed intervention (“turn down study”) and (B) for pharmacologic intervention at 3000 RPM.

Also indicated in the figure for reference is the recovery criterion reported in RESTAGE of EF as 45%, confirming partial recovery at POD 360. TAVEF on the other hand does not exceed this value at 3000 RPM. This suggests that a speed-dependent threshold be introduced for evaluating recovery based on TAVEF, as illustrated in Fig. 5A.

Intuitively the degree to which LVEF responds to changes in VAD Speed depends on the LV Preload at the baseline speed. It is expected that for NYHA I or patients with greater myocardial recoverty, both TAVEF and LVEF be more sensitized to change with decreasing VAD Speed depending on the change in LV Preload. Yet, the results indicate only TAVEF to show significant change to VAD turn-down. This indicates an important point on the interaction between the Heart and the VAD leading to the dyanmics of the cooperative mechanism, that in the dialted ventricular model the study employs there is an observed insensitivity in LVEF for the VAD Speed.

The chronically pathologically overloaded LV benefits from mechanical unloading into a range of normal loading conditions; the acutely pathologically overloaded LV, by contrast, likely benefits from more aggressive unloading, particularly in the setting of myocardial ischemia. Reducing loads to a subnormal level may benefit myocardial energetics, but likely is otherwise injurious/cytotoxic to cardiomyocytes. Usually, things that are ought to move within a physiologically normal range, as this is part of muscle homeostasis. Not exercising muscles is a bad thing for them, and the heart is not different. Some level of preload is good for the muscle, while some level of afterload probably is important for myocardial perfusion.

This study considered spectrum of heart failure from normal/healthy to NYHA IV, by employing lumped-parameter values derived from clinical hemodynamic data, by Ky et. al. Nevertheless, by being an in-silico study, it is limited, and therefore demands clinical validation. For example, at 0 RPM, the simulation counterintuitively predicts SVVAD to approach zero, but does not become significantly negative. This is because of inertance in the outflow graft in the simulation. Further studies are required to dwell deeper into this subject, to have a complete understanding of the effect of inertance on retrograde flow in the VAD, using in-vitro experiments. Anecdotal reports corroborate this observation, however clinical validation would provide an important guideline for future turn-down studies. A second limitation is that there is currently no real-time method to measure TAVEF but requires use of echocardiography or an equivalent way to measure aortic ejection. Therefore, development of a non-invasive measure of aortic flow would be a valuable contribution. Studies pertaining to the model classification, in terms of the unimodal ESPVR as compared to the linear conventional ESPVR towards the sensitivity of TAVEF and LVEF need to be undertaken to gather a more holistic picture, that also would help understand the interaction between the Heart and the VAD in a recovering patient.

## CONCLUSION

The aim of the study was to understand and lay the foundation to evaluate ventricular recovery through the interplay between the native ventricle and the VAD, specifically focusing on the opening of the aortic valve. Trans aortic valve ejection fraction, TAVEF, was introduced to help understand this dynamic interaction and provide a more sensitive metric for preload-sensitive response of the native ventricle. Based on these encouraging results, future clinical investigation is warranted to evaluate the efficacy of TAVEF as a measure of ventricular recovery.

## Data Availability

No additional data is available and all the data used in the study are included in the manuscript and supplemental publication materials.

## Abbreviations

AoV: Aortic Valve
AoVC: Persistent Aortic Valve Closure
BTR: Bridge to Recovery
CO: Cardiac Output (lpm)
CVS: Cardiovascular System
DBP: Diastolic Blood Pressure (mmHg)
DP: Dopamine
E_max_: Maximal Elastance
EDV: End-diastolic Volume (ml)
ESP: End-Systolic pressure in mmHg
ESV: End-Systolic Volume (ml)
LVAD: Left Ventricular Assist Device
LVEF: Left Ventricular Ejection Fraction
MAP: Mean Arterial Pressure (mmHg)
MCS: Mechanical Circulatory Support
NYHA: New York Heart Association
RPM: Rotations Per Minute
SBP: Systolic Blood Pressure (mmHg)
SNP: Sodium Nitroprusside
SoS: System of Systems
SV: Stroke Volume (ml) (=EDV – ESV)
TBV: Total Blood Volume
TAVEF: Trans-Aortic Valvular Ejection Fraction
TAVF: Trans-Aortic Valvular Flow
TAVO: Trans-Aortic Valvular Output
TVF: Trans-VAD Flow
VAD: Ventricular Assist Device
VBE: Volume in Left Ventricle Before Ejection Phase

## ACKNOWLEDGMENT

This work was supported by and performed by contractors of the US Government Grant, W81XWH2010387.

## Notes

### Competing Interest Statement

The authors have declared no competing interest.

### Author Declarations

No clinical study was done for this manuscript and hence no IRB protocols have been included.

